# Mortality Improvements in Adult Patients Hospitalised with Community Acquired COVID-19 in Wales From March 2020 to December 2021

**DOI:** 10.1101/2022.08.26.22279219

**Authors:** Simon M Barry, Gareth R Davies, Chris R Davies, Keir E Lewis

## Abstract

**Background:** A COVID-19 hospital guideline was implemented across all acute hospitals in Wales in March 2020, and data was collected across the first 3 Waves of the pandemic. We aimed to observe trends in mortality with a focus on ward-based outcomes.

**Methods:** Retrospective case-note review of data for adults admitted to hospital with community acquired COVID-19 between March 2020 and December 2021

**Results:** 5887 cases were analysed. Overall mortality from COVID-19 fell from 31.5% in Wave 1 to 22.6% in Wave 2 to 18.8% in Wave 3 (p<0.01). Ward mortality for patients on oxygen fell from 34.6% in Wave 1 to 19.5% in Wave 2 (p<0.01) to 14.3% in Wave 3 (p=0.03). For those managed with CPAP/HFNO on wards, the mortality reduced from 58.9% in Wave 1 to 45.6% in Wave 2 (p=0.05) and further to 42.6% in Wave 3 (p=0.03). The mortality for patients managed with CPAP/HFNO on ICU reduced from 43.8% in Wave 1 to 24.7% in Wave 2 (p=0.12) and further to 20.4% in Wave 3 (p=0.03). Patients receiving CPAP/HFNO on the wards were on average 11 years older and more co-morbid than those on ICU. In Wave 3, 77% of hospital admissions with COVID-19 were unvaccinated with mortality rates of 20.5% compared to 4.8% mortality in those who had received three vaccines (p<0.01).

**Conclusions:** There were successive reductions in mortality in inpatients over the 3 Waves reflecting new treatments and better management of complications. The impact of vaccines on outcomes of hospitalised patients was notable in Wave 3.

**Key Messages:** 

**What is the key question?:** What are the outcomes from COVID-19 pneumonitis managed on respiratory wards and how have they changed over successive waves of the pandemic?

**What is the bottom line?:** Significant improvements in mortality over time were noted in patients requiring oxygen, CPAP or HFNO. Patients managed with these modalities in ICU had lower mortality rates than those on wards, but they were younger and less co-morbid. In wave 3 patients were largely unvaccinated with higher mortality rates than those who were fully vaccinated.

**Why read on?:** This is a national study including all acute hospitals in Wales over three waves of the pandemic from March 2020 to December 2021. It is the first paper to demonstrate at a national level the outcomes of ward management of COVID pneumonitis over successive waves.

## INTRODUCTION

As of July 2022, the number of reported cases of COVID-19 exceeds 570 million worldwide, with more than 6.3 million deaths,^1^ although the true figure is likely to be much higher due to underreporting.^2^ In Wales, a country with a population of 3.2 million people with health care delivered by seven regional Health Boards (HBs) in a devolved National Health Service there have been 880,000 cases and 7605 deaths.^3^ In March 2020, as the pandemic unfolded, and hospital admissions rose across the UK, in the absence of UK guidance Wales launched its national COVID-19 hospital guideline. The guideline was developed and disseminated through a digital implementation framework, Simple IMPlementation ScIence (SIMPSI),^4^ deploying local facilitators to maximise guideline adoption, particularly targeting senior clinical decision makers (Consultants). The guideline was hosted on a digital platform requiring user registration and updates were delivered to healthcare registrants in a video format from experts in the practical management of COVID-19. In its first 3 months, the platform rapidly gained 4521 registrants including the vast majority of senior clinicians in frontline specialities delivering acute COVID care, achieving 170,000 views.^5^ An early consensus decision was taken in Wales to manage patients with hypoxaemic COVID pneumonitis on respiratory wards and avoid invasive ventilation where possible, and this decision was incorporated into our COVID hospital guideline. In addition, Welsh Government approved a national data collection tool, initially to collect data from Wave 1 but subsequently expanded to include Waves 2 and 3. Therefore, Wales developed an early strategy to try and standardise acute hospital care promoting ward management of COVID pneumonitis and to systematically collect data on the outcomes of the COVID-19 pandemic at a national level.

In the first national report on outcomes from COVID-19 during Wave 1, we found increased death rates in those who contracted COVID-19 in hospitals (nosocomial acquired) compared with those who were admitted with community acquired COVID-19, reflecting the increased frailty and co-morbidities seen in patients who were already in hospital.^6^

We now report outcomes from community acquired COVID in hospitalised patients across three distinct Waves of the pandemic. We examine the impact of age, co-morbidities, and deprivation index on mortality at each Wave, as emerging therapies, medical experience and vaccination affected outcomes over time. We report outcomes from respiratory wards and ICUs.

## METHODS

Retrospective observational study of prospectively gathered data of adults (aged over 18 years) treated in hospital for at least 24 hours in all 18 acute hospitals in Wales.

### Study design

A digital tool collected anonymised patient and site-level outcomes (www.audit.clinicalscience.org.uk). This tool was endorsed by Welsh Government and the Information Governance Departments in each HB. It was designed and implemented through the Institute for Clinical Science and Technology (ICST), who identified hospital audit and clinical leads to manage local data collection. The data collection tool was hosted within the National Pathway for Managing COVID-19 Infections in Secondary Care in Wales initiative (www.allwales.icst.org.uk) which also hosts the COVID-19 hospital guideline.

Denominator data obtained from Public Health Wales (PHW) was all patients over 18 years admitted to hospital with a positive SARS-CoV-2 polymerase chain reaction (PCR) result between 1^st^ March 2020 and 14^th^ December 2021[5]. This data was cross-checked with local hospital admission records to identify patients for continuous notes review by local clinical teams. Community acquired COVID-19 was defined as symptoms developing in the community and a positive PCR test in the community up to two weeks before admission, or within 48 hours of admission.

### Data collection

Demographic variables were collected for the index admission and inputted into the online tool. Mandatory fields included date of positive PCR swab, date and site of admission and discharge, age, gender, and outcome (death or discharge). Vaccination status was also a mandatory field for Wave 3. Supplementary fields included obesity, number of co-morbidities, frailty score and type and location of treatments given. Where patients received ICU and ward-based care or CPAP and ‘oxygen alone’, we allocated them to the highest-level-of-treatment group. The Welsh Index of Multiple Deprivation (WIMD) was derived from the patient’s post-code and is the Welsh Government’s official measure of relative deprivation for small areas in Wales.^7^ Available hospital notes were retrieved by record departments from the denominator lists.

Wave 1 was between March 1^st^ 2020 to November 1^st^ 2020

Wave 2 was between November 2^st^ 2020 and February 21^st^ 2021

Wave 3 was between June 1^st^ 2021 and December 14^th^ 2021

These 3 Waves were defined as they represent distinct spikes in the population prevalence of SARS COV-2 variants (figure 1), relating predominantly to the wild type (wave 1), alpha (wave 2) and delta (wave 3) variants.^8^ We stopped data collection in mid-December at the onset of the omicron wave.

**Figure 1.**
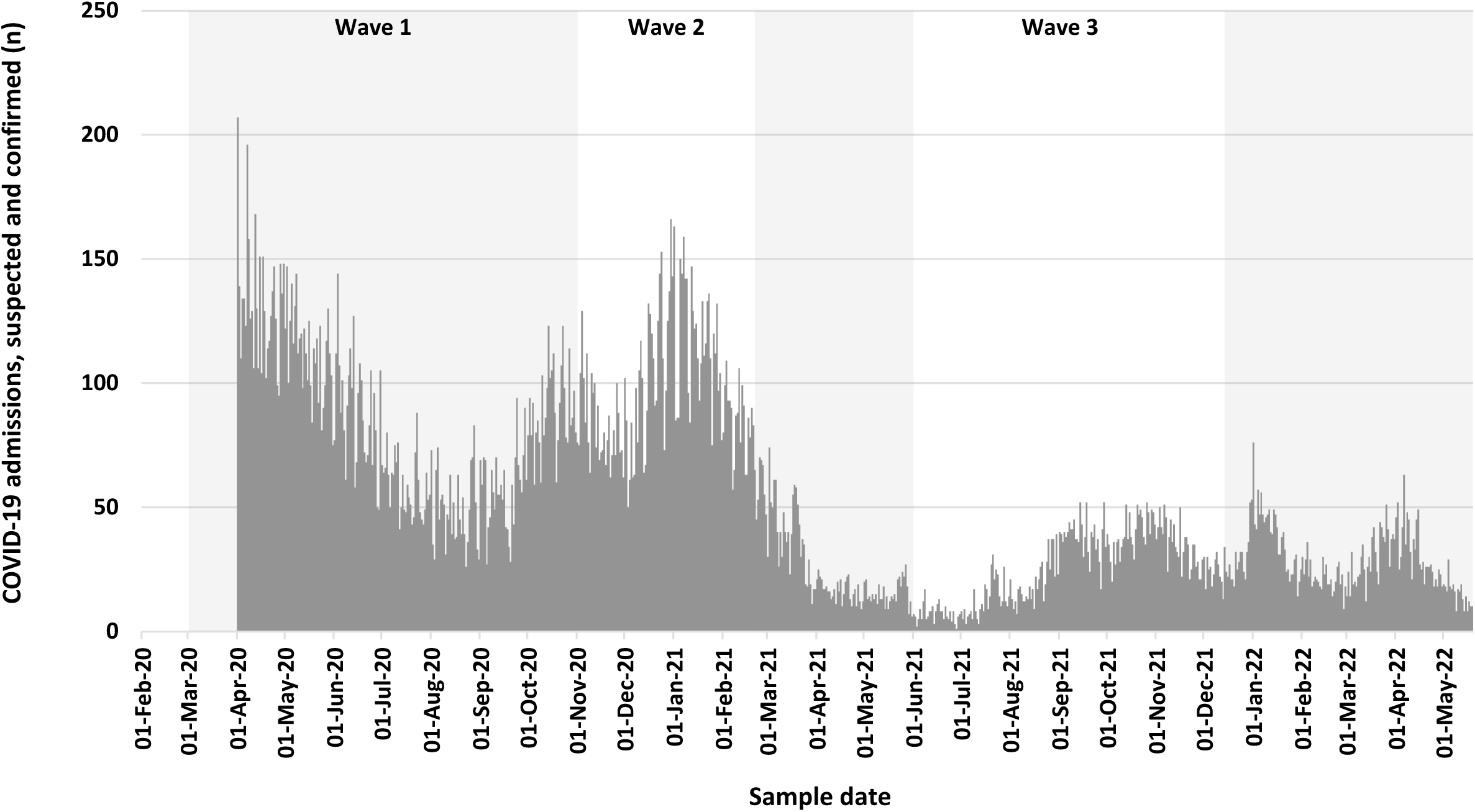
Number of confirmed COVID-19 hospitalisations in Wales aged over 18 from March 2020 to December 2021, corresponding to Waves 1-3.^9^

### Outcomes

The primary outcome was mortality, which was assessed across the Waves univariately by age, gender, comorbidity, admission setting (ward or ICU), deprivation and modality of respiratory support. We also considered mortality by vaccination status for Wave 3.

### Missing data

Data were tabulated as the percentage completed for each parameter by HB in each Wave. Mandatory fields achieved 100% completion but there was a large variation by HB for some of the non-mandatory fields (Table 1). Data completion for WIMD was 95% since some post codes did not link to a deprivation status.

**Table 1.**
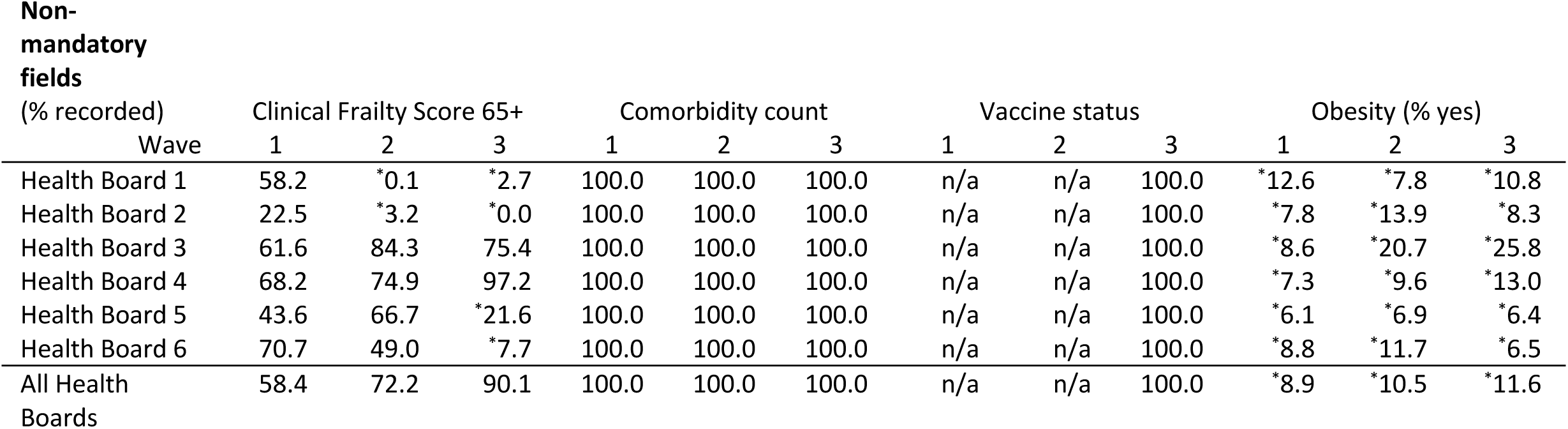
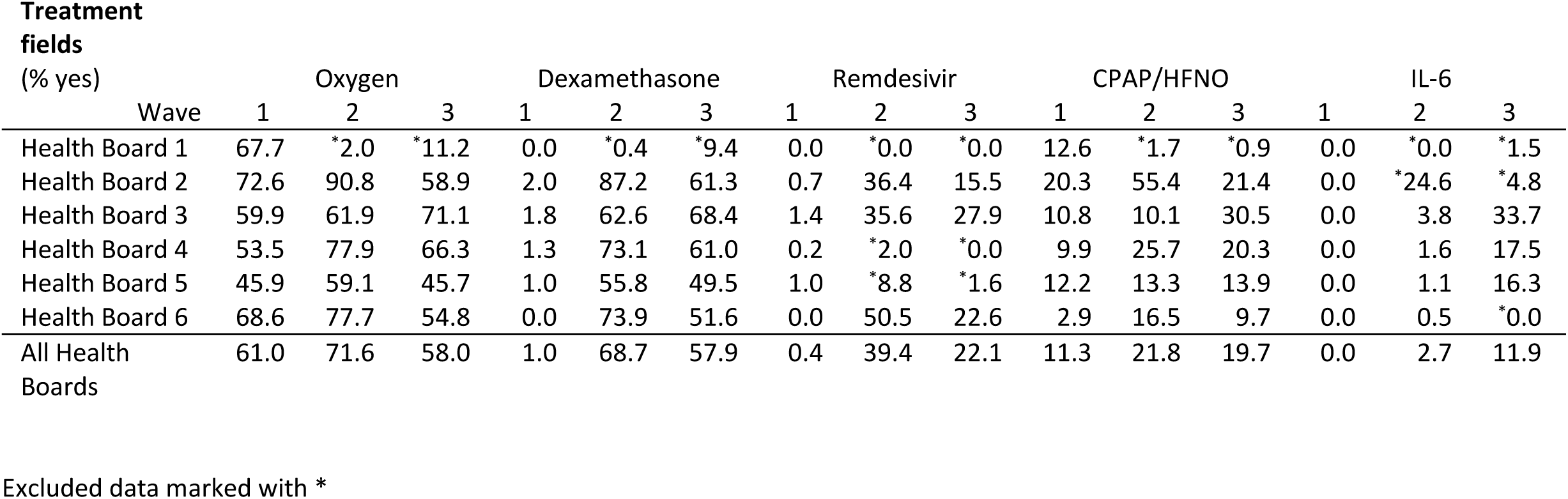
Percentage completion for non-mandatory fields across all three Waves by Health Board.

Parameters were excluded from analysis if deemed to be outliers for no obvious clinical reason (e.g. CPAP was not delivered in that hospital). No missing data imputation was used. BMI was rarely recorded in the notes and so levels of recorded obesity do not reflect true prevalence rates of 33%.^10^

### Statistical methods

Baseline demographic and clinical characteristics and outcomes were combined across the Health Boards (18 hospitals) and were grouped by Wave. Continuous variables are presented as median [inter-quartile range, IQR] and categorical variables as n (%), unless otherwise stated.

Univariate analysis of categorical data used either Fisher’s exact test or paired proportions test. For continuous data, t-tests and Mann-Whitney U tests were employed. Cochran-Armitage tests for trend were used to evaluate rates across the Waves. Two-sided statistical significance was set at p=0.05 for all tests.

A multivariable logistic regression model was used to assess the effect on mortality of each covariate after adjusting for all the other covariates, namely: Wave, total comorbidities, 10-year age band, gender and deprivation grouping. The largest category in each covariate was used as the baseline category. No attempt was made to reduce this saturated model and no interaction terms were considered. No validation of the model on a subset or additional data set was undertaken.

The saturated model was assessed by means of the Pearson Chi-square goodness of fit test and the Area under the ROC curve. Residual plots were inspected for outliers. Outlying covariate patterns were assessed by the project team. All analyses were conducted in Stata BE 17.0.

## RESULTS

For Wave 1, we had datasets from 1563 patients, for Wave 2 there were 3039 patients and for Wave 3 there were 1842 patients. Of these 6444 datasets, we removed 179 duplicates, 298 patients with length of stay less than 24 hours, and 80 cases under 18 years old, leaving a total of 5887 patient datasets for analysis. This represents 40% of all admitted cases (n= 14,614 across the three waves).

Overall there were 1398 inpatient deaths from the 5887 patient data sets (mortality rate 23.8%).

The age of admission was similar for Wave 1 and 2 but those admitted in Wave 3 were 4 years younger (p<0.01) There was no difference in the gender or proportion from the most deprived 30% over the 3 Waves (table 2).

**Table 2.** Sociodemographics of hospitalised COVID patients across the 3 Waves

| **Wave** | Median Age (IQR) | % Male | % from most deprived 30% |
| --- | --- | --- | --- |
| 1 | 67.3 (56.5 to 80.0) | 56.9 | 41.0 |
| 2 | 66.7 (55.0 to 80.0) | 54.4 | 41.3 |
| 3 | 62.9 (49.0 to 78.0) | 54.7 | 41.5 |
| p value | <0.01 | 0.27 | 0.97 |

Univariate logistic regression analyses produced significant odds ratios for all variables considered except deprivation. There odds ratios for wave 1 vs 2 and 2 vs 3 showed significant reduction in mortality from wave to wave. The odds ratios for 0 to 5+ comorbidities exhibited a clear gradient of increasing mortality with increasing comorbidities. The effect of age was not so linear but showed significantly lower odds ratios in all ages before 70-79 and significantly higher odds ratios for ages from 80 onwards. The odds ratio for sex indicated significantly lower mortality in females. There were no statistically significant odds ratios for any of the deprivation groups when compared to the 50% least deprived (figure 2).

**Figure 2.**
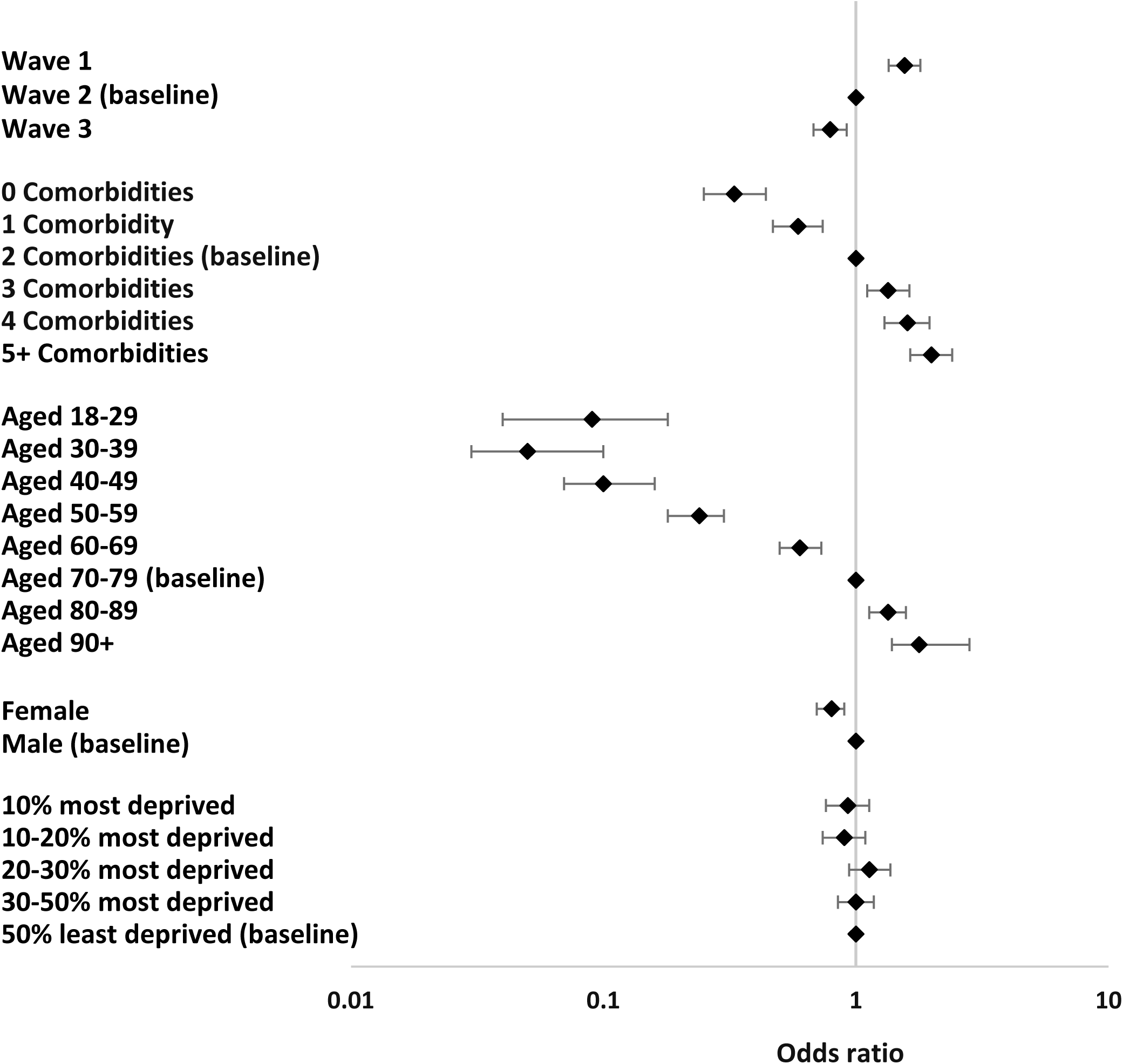
Univariate odds ratios for mortality by wave, co-morbidities, age, sex and deprivation

Adjusting for all the covariates simultaneously in a multivariable logistic regression model made no substantive difference to the conclusions reached from the univariate analyses. The effect of age persisted with lower odds at lower ages and greater odds at older ages, with the odds ratios for the oldest ages increasing slightly compared to the univariate analysis though the difference between waves 2 and 3 narrowed. The positive relationship between increasing comorbidities and increasing mortality remained, though there was no substantive difference between 2 & 3 comorbidities now. Females retained the significantly lower odds of mortality over males and there were no statistically significant differences between deprivation groups, though the most 20-30% deprived was almost higher than the least 50% group (p=0.05), figure 3.

**Figure 3.**
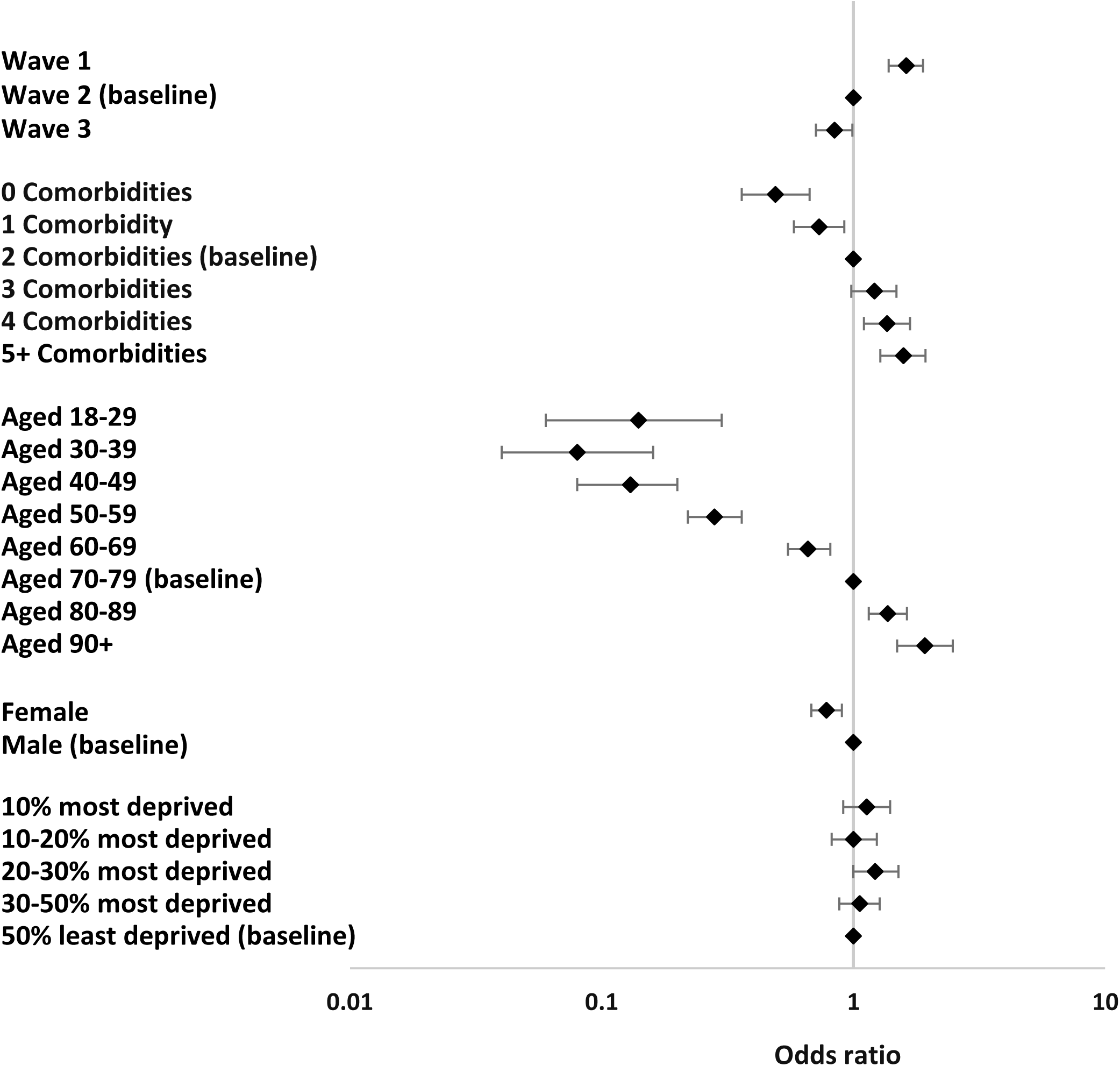
Multivariate odds ratios for mortality by wave, co-morbidities, age, sex and deprivation.

The Pearson Chi-square goodness of fit test did not indicate any problems with the model fit (p=0.41) and the area under the ROC curve was 0.75 which is within the range deemed acceptable in clinical discrimination models (0.70-0.79). Regression diagnostics (change in deviance, Chi square and Cooks distance) indicated 50 outlying patients’ covariate patterns totaling 252 observations. Upon inspection these patterns were generally of combinations of higher clinical risk factors (e.g. 83 years old with 5 co-morbidities) with little or no resultant mortality. None were deemed clinically unreasonable, so all records were retained in the model.

Mortality rates and admission numbers for all patients was compared with numbers and admissions to ICU. HB 1 was excluded due to incomplete data, leaving 3785 for analysis. The combined all-cause mortality declined from 31.6% (Wave 1) to 22.0% (Wave 2) to 18.6% (Wave 3) (p<0.01, Cochran-Armitage test for trend), Figure 4.

**Figure 4.**
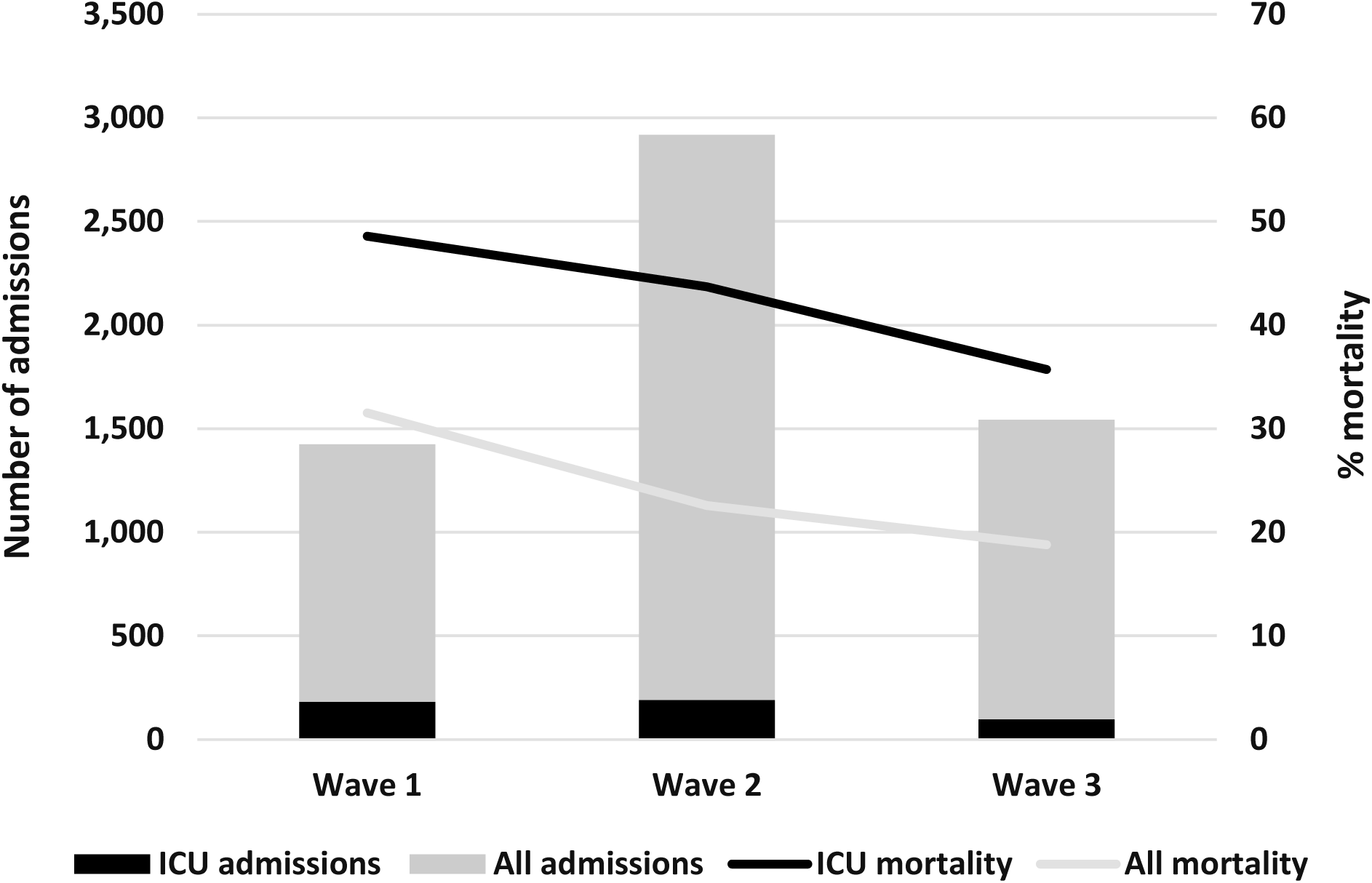
Mortality for all patients and ICU patients Waves 1-3.

The proportion of admitted patients who were managed in ICU (at least at some point) remained relatively constant at 11.6%, 10.7% and 9.3% (p=0.10 Cochran-Armitage test for trend). ICU mortality reduced from 48.6% (Wave 1) to 43.7% (Wave 2) to 35.7% (Wave 3) (p=0.02, Cochran-Armitage test for trend).

For patients who survived to discharge, the median length of stay in Wave 1 and 2 was 7 days (IQR 3-17) but reduced to 5 days (2-11) for Wave 3 (p<0.01 Wilcoxon rank sum test). For patients who were discharged from ICU, the median length of stay was 29 days (16-47) in Wave 1, 18 days (10-35) in Wave 2 (p<0.01) and 15 days (10-25) in Wave 3 (p=0.25 for Wave 2 vs Wave 3).

Next, we analysed mortality rates of only ward-based patients according to respiratory support (n=3785, figure 5). This analysis also excludes HB 1, where there were incomplete data for these parameters (see Table 1). There was a significant reduction in the mortality of patients managed on wards with oxygen alone from 34.6% in Wave 1 to 19.5% in Wave 2 (−15.0%, 95%CI: 9.8 to 20.2, p<0.01) and a further smaller drop to 14.3% in Wave 3 (−5.2%, -0.7 to -9.7, p=0.03). There was a reduction in mortality of patients managed on wards with CPAP/HFNO from 58.9% in Wave 1 to 45.6% in Wave 2 (−13.3%, -0.2 to -26.4, p=0.05) and further smaller drop to 42.6% in Wave 3 (−5.2%, - 0.7 to -9.7, p=0.03).

**Figure 5.**
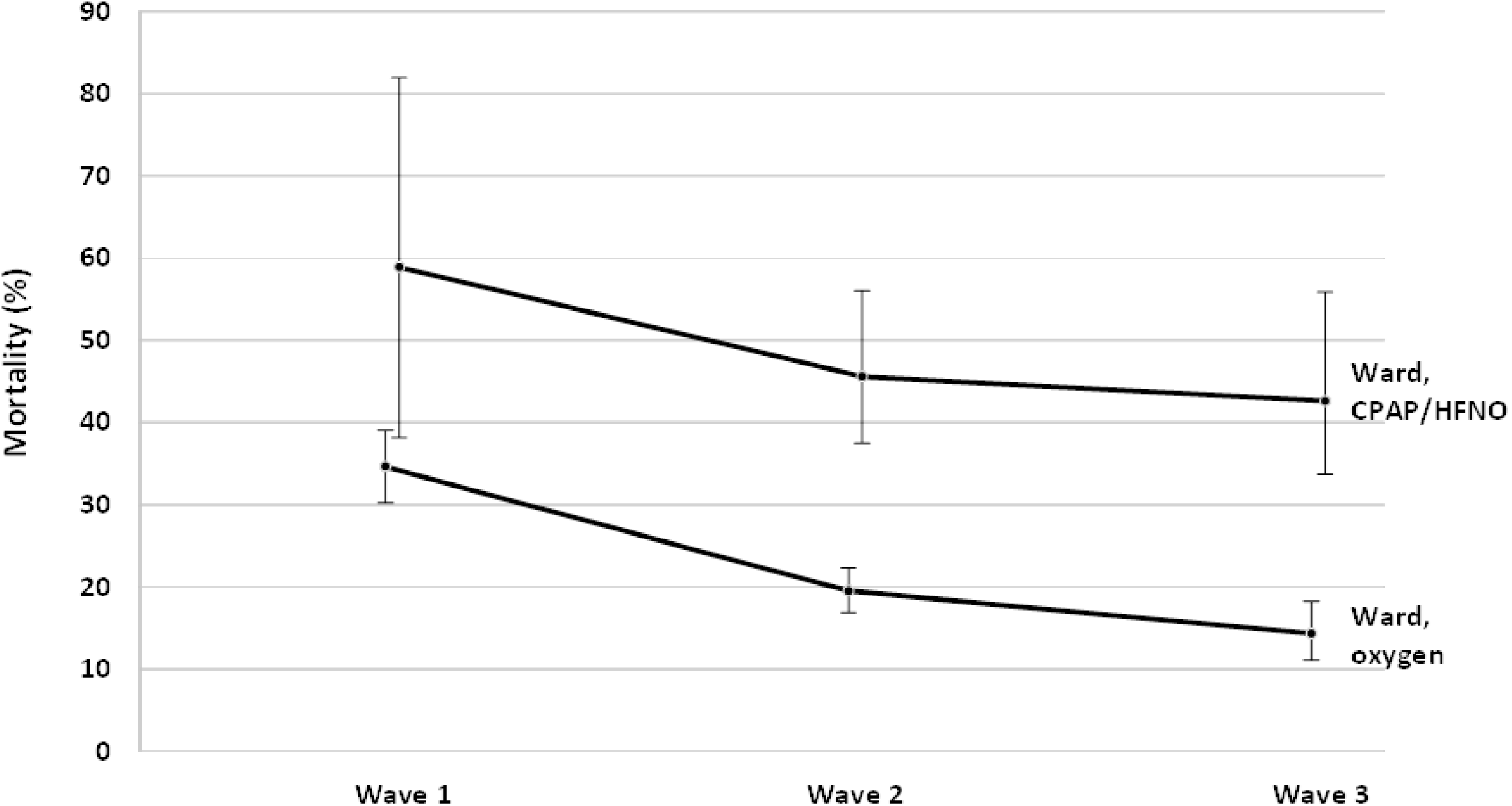
Mortality rates by treatment modality on wards

The mortality of patients managed with CPAP/HFNO in ICU was 43.8% in Wave 1 and 24.7% in Wave 2 (−19.1%, -7.0 to +45.1, p=0.12) and 20.4% in Wave 3 (−4.3%, -10.4 to -19.0, p=0.03).

Combining all 3 Waves, those patients receiving CPAP/HFNO on the wards were older (median 69 years, IQR 58-77) than those receiving CPAP/HFNO on ICU (median 58 years, IQR 50-65 years), p<0.01 and this difference of 10-11 years was similar in each individual Wave. Across all 3 Waves combined, patients receiving CPAP/HFNO on the wards had more comorbidities (median 3.0, IQR 2.0 to 4.0) than those patients receiving CPAP/HFNO in ICU (median 2.0, IQR 1.0 to 4.0) (p=0.01).

We next analysed mortality by wave, age and deprivation, dividing the cohort into the most deprived 30%, then next most deprived 20% and the least deprived 50% for visual clarity (figure 6). As expected, mortality increased with age, particularly in those older than 70 years, but most striking was the reduction in mortality for all levels of deprivation between successive Waves in this more elderly group.

**Figure 6.**
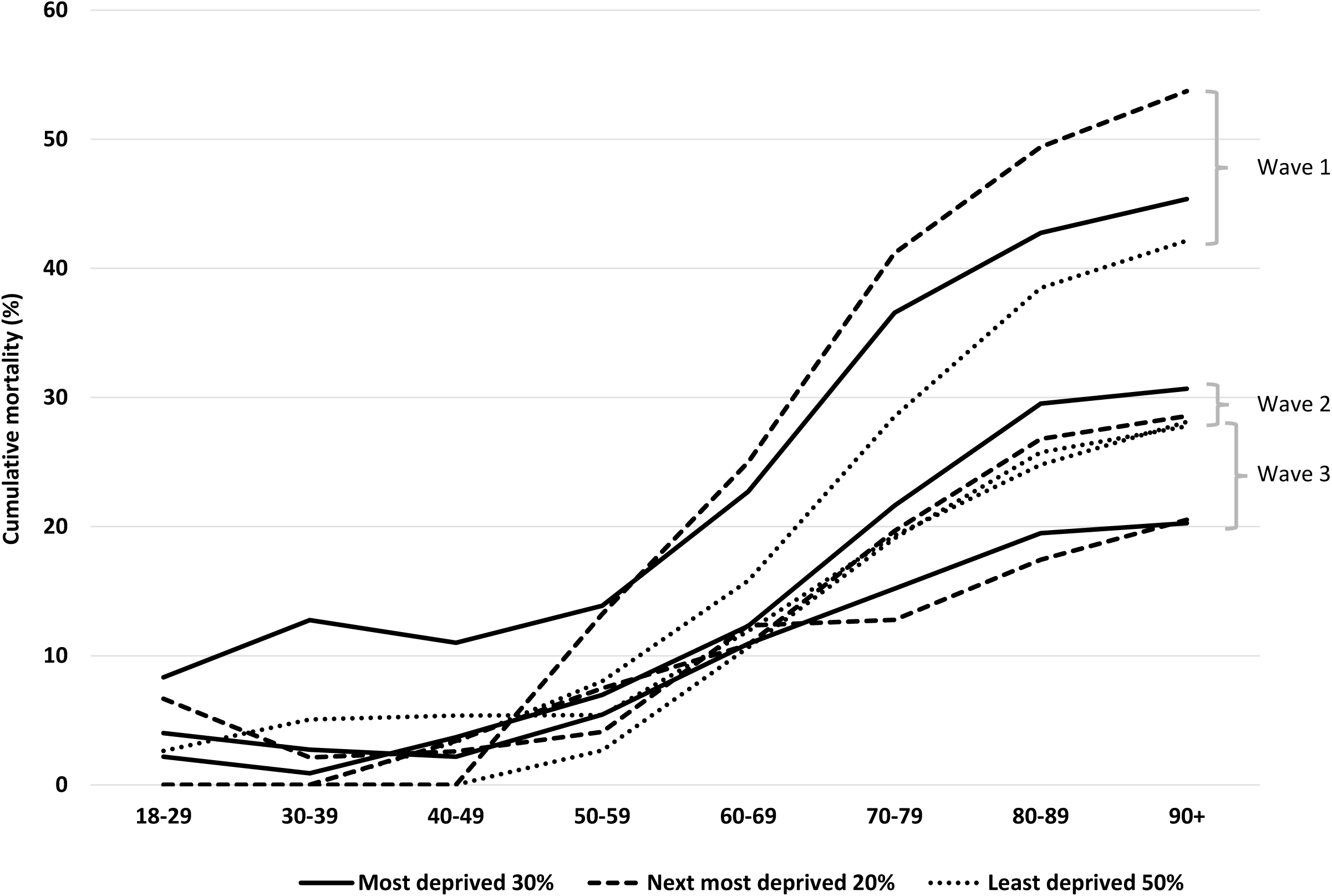
Mortality by age and deprivation for each Wave

For Wave 3, 77% of people admitted with COVID were unvaccinated, compared to 5% admissions who had full vaccination (at that time two vaccines and a booster). The overall mortality was 20.5% of those with no vaccine cover, compared to 4.8% of those fully vaccinated (p<0.01). In Wave 3, 81% of people from the most deprived 10% were unvaccinated, compared to 73.4% in the ‘least deprived 50%’ being fully vaccinated (p=0.03).

## DISCUSSION

We present sociodemographic data and outcomes of patients admitted to every acute hospital in Wales, with community acquired COVID-19 through 3 distinct Waves over the first 21 months of the pandemic. This National Report is the culmination of a strategic attempt to standardise hospital care for patients in Wales with COVID-19 using a unique digital management guideline and to collect data on outcomes.

The data on 5887 admissions represents 40% of total community acquired COVID-19 admissions in acute hospitals in Wales. Our overall mortality rate of 23.8% was consistent with other studies,^11-13^ but higher than the 5% reported in Japan.^14^ This latter study comprised a younger cohort with 3% under 18, 22% aged 18-39 and 41% over 65. Our patients were considerably older with only 9% in the 18-39 age group, 57% aged over 65 and we excluded those under 18. In addition, only 26% of the Japanese patients required oxygen and 2.8% HFNO/CPAP, compared to 64% on oxygen and 17.6% on CPAP/HFNO on wards in Wales, suggesting a cohort with less severe pneumonitis. Our patients had higher mortality with increasing co-morbidities, age and male sex, consistent with previous studies.^11-14^

The mortality rates for those in ICU and anyone on CPAP/HFNO were higher than those with oxygen alone at each Wave, indicating a group with more severe pneumonitis. Our longitudinal analyses found mortality improvements for each treatment group across successive Waves. The reduction in all-cause mortality over time reflects treatment effects and it is notable that the biggest reduction was between Wave 1 and Wave 2 where the introduction of dexamethasone became the standard of care for anyone requiring oxygen.^15^ There were improvements in other ward-based treatments between Waves 1-2 for example a better balance between CPAP and HFNO for those who required more prolonged treatment, more extensive use of proning, a lower threshold for testing for pulmonary emboli and awareness of complications such as pneumomediastinum and superadded infections. These and other management strategies were disseminated to registrants via updates on the Wales COVID hospital guideline. By end of Wave 2 there were further impacts of IL6-inhibitors^16^ and more experience with CPAP and by Wave 3, vaccines^17,18^ and anti-COVID antibody therapies^19^ were impacting on the severity of illness.

We found that those in the ‘most deprived 30%’ were over-represented, accounting for 42% of hospital admissions with COVID-19. There was an improvement in mortality for all levels of deprivation with each successive Wave suggesting all patients had similar access to improvements in hospital medical care over time. Multivariate analysis indicated that deprivation was not an independent risk factor for mortality overall. This finding is at variance with other data^20,21^ and merits further discussion. Our analysis is only of hospitalised community acquired COVID-19 in adults so cannot reflect population level associations between COVID mortality and deprivation. Indeed, Office for National Statistics (ONS) data demonstrated a clear association between deprivation and mortality in England and Wales.^22^ Our findings of increased admissions from the most deprived 30% suggest that access to healthcare was not a major factor and the lack of effect of deprivation on mortality in hospital is reassuring.

Unvaccinated patients represented 77% of admissions during Wave 3 and the unvaccinated had a mortality rate of 20.5% compared to 4.8% in those double vaccinated with a booster. This confirms the strong protective effects of vaccines against hospitalisation and death.

There are several strengths to our study: First, this was a truly national dataset from all acute hospitals in Wales on almost half of all COVID admissions. Second, due to the online data collection tool, there were complete datasets for mandatory fields. Third, we applied a strict definition of community acquired COVID with prior symptoms and a positive test within 48 hours of admission, thus avoiding the inclusion of nosocomial cases that are a distinct sub-group with a higher mortality rate.^6^ Fourth, our hospital COVID-19 guideline was widely disseminated and the first to measure user activity thus supporting standardisation of care in Wales. At the beginning of the pandemic, key individuals in the respiratory and intensive care community agreed firstly that ‘permissive hypoxaemia’ should be allowed, and the target oxygen saturations in Wales were set at 90-94%, lower than the 92-96% recommended in England.^23^ Consensus was also reached early in wave 1 that patients should be managed on respiratory wards and given oxygen, with the addition of CPAP or HFNO according to the guideline and only those failing on this treatment who were suitable for escalation, or who initially presented with severe hypoxaemia should be managed in critical care. This decision was taken partly in order to protect limited ICU beds as Wales has a lower number per capita than England and about half that of the European average,^24^ but also because there was a consensus view early on that invasive ventilation was best avoided. It is of interest therefore that UK wide ICU data collection demonstrated that Wales had a lower proportion of patients managed in intensive care on basic respiratory support (oxygen, CPAP and HFNO) compared to the UK as a whole (16.4% compared to 25.7% for the first wave and 27.3% compared to 42.8% for the second wave),^25^ suggesting that the Welsh national approach was followed with this support occurring predominantly outside of the intensive care setting on respiratory wards.

Our national experience thus provides evidence supporting ward management of hypoxaemic COVID-19 pneumonitis with continued improvements over time. This data supports the adoption of Respiratory Support Units to manage patients with respiratory failure.

This study suffers from the limitations of retrospective case-note reviews with missing data on non-mandatory fields including frailty scores and treatments particularly from one HB (table 1). Obesity, a recognised risk factor for COVID^26^ was also poorly recorded. Our prerequisite for symptoms and positive swab within 48 hours of admission would have missed a proportion of community acquired cases that could have had a positive PCR up to 5-7 days after admission, but we have argued this approach could have included nosocomial cases. We do not have data relating to severity of co-morbidities (see ICNARC) and clinical severity on admission. We have not mapped the outcomes against variants of the virus, but the Waves largely corresponded to the Wuhan wild type, alpha and delta waves respectively. Finally, whilst we have collected data on a large proportion of admissions across all acute Hospitals in Wales, we cannot exclude case ascertainment bias according to notes availability although with over 5800 records, we feel this is unlikely.

We provide national data from 18 hospitals over 21 months, following the implementation of a hospital guideline supporting the ward management of patients with COVID-19 pneumonitis. We report declining mortality rates with each successive wave highlighting the beneficial effects of new treatments and vaccination. Our study provides insights for dealing with future pandemics particularly focussing on the implementation of new evidence as it emerges to a target audience.

## Data Availability

all data produced in the present work are contained in the manuscript

## Acknowledgements

Public Health Wales for providing the denominator data on COVID admissions, Hannah Sharp (Institute Clinical Science and Technology) and the local facilitators Sarah Bowen, Carol Llewellyn-Jones, Abdelrahman Mohamed, Inder Singh, Shehnoor Tarique, Carla Dos Santos Gil, Sara Fairbairn, Tarek Dihan, Rhodri Edwards, Matt Brouns, Ramsey Sabit, Amit Benjamin, Jacqueline Woolley, Claire Kilduff, Daniel Menzies, Liz Brohan, Alexandra Scott, Sinan Eccles, Anna Lewis, Sharon Ragbetli, Sion O’Keefe, Joanne Stimpson, Laura Gingell, Liz Evans, Ian Bebb, Jane Christmas and Favas Thaivalappil. All junior doctors and audit teams who helped input data.

## Contributors

SB conceived the COVID-19 hospital guideline and database and wrote the first draft of the paper with KL. GD performed all of the statistical analysis and CD created and implemented the COVID-19 digital guideline and database. All authors critically revised and approved the final version.

## Patient consent for publication

Not required

## Ethics approval

This work was performed as a service evaluation. A formal waiver for ethical approval was obtained from Approvals Officer Dr Beresford (Welsh REC2). All Health Board information governance leads approved the digital data collection tool that did not collect patient identifiable information

